# Is tracking and modeling Covid-19 infection dynamics for Bangladesh using daily data feasible?

**DOI:** 10.1101/2020.06.13.20130617

**Authors:** Karar Zunaid Ahsan, Rashida E Ijdi, Peter Kim Streatfield

## Abstract

Given the low Covid-19 testing coverage in the country, this study tested whether the daily change in the number of new Covid-19 cases is due to increase (or decrease) in the number of tests done daily. We performed Granger causality test based on vector autoregressive models on Bangladesh’s case and test numbers between 8 March and 5 June 2020, using publicly available data. The test results show that the daily number of tests Granger-cause the number of new cases (p <0.001), meaning the daily number of new cases is perhaps due to an increase in test capacity rather than a change in the infection rates. From the results of this test we can infer that if the number of daily tests does not increase substantially, data on new infections will not give much information for understanding covid-19 infection dynamics in Bangladesh.

## Introduction

First reported in Wuhan, the capital city of Hubei province in Central China, in December 2019 (Li et al., 2020), 2019 novel coronavirus disease (COVID-19) now poses an unprecedented challenge for Bangladesh and many other developing countries. Between 08 March and 12 June, 2020, the total number of persons with laboratory confirmation of Covid-19 infection in Bangladesh increased from 3 to 68,504—putting Bangladesh among the highest 20 countries in the world in terms of total number of Covid-19 cases.

Based on daily data on new cases and tests done, as reported by the country’s official Covid-19 information agency the Institute of Epidemiology, Disease Control and Research (IEDCR) and reported in the official dashboard (GOB, 2020a), many researchers in Bangladesh and abroad are analyzing patterns in infection rates and projecting daily case numbers to examine the infection dynamics of Covid-19 in Bangladesh. As the collection, dissemination and use of good data on testing is a key part of the global response to the pandemic (WHO, 2020), such effort is greatly beneficial for public health research and crucial for health systems planning to address this ongoing crisis.

Until recently, the number of tests per capita in Bangladesh was among the bottom ten countries for number of tests. From the very beginning the number of daily new cases and the number of daily tests is closely related (correlation coefficient 0.98) (Khan 2020). Though strong correlation does not necessarily imply causation, such a strong, positive relationship between new case and daily test numbers that has been consistently going on for the last three months raises a question: Can the daily change in the number of new Covid-19 cases be just due to increase (or decrease) in the number of tests done daily? Until now, no specific causality analysis of the mutual relationship between these two variables has been conducted for Bangladesh.

## Data and Methods

We used daily Covid-19 reports for Bangladesh between 08 March and 05 June, 2020 from Our World In Data (Roser et al., 2020) to test for Granger causality between two variables: daily new cases and the number of tests done in the past 24 hours. Granger causality is based on vector autoregressive (VAR) model, where a two variable VAR will look like:

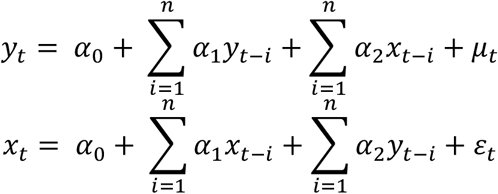

where *y*_*t*_ is daily new laboratory confirmed cases, where *x*_*t*_ is daily tests done, *μ*_*t*_ and *ε*_*t*_ are white noise error processes, and n denotes the number of lagged variables. Variable *x*_*t*_ is said to “Granger-cause” variable *y*_*t*_if, given the lags of *y*_*t*_, the lags of *x*_*t*_ are jointly statistically significant in the *y*_*t*_ equation. All the analyses were performed in Stata 16.

## Results and Discussion

During the 90-days study period, we found a near-perfect relationship (*r* = 0.97, *p* < 0.001) between daily new cases and tests done, and the relationship is statistically significant (see Figure 1).

**Figure 1.**
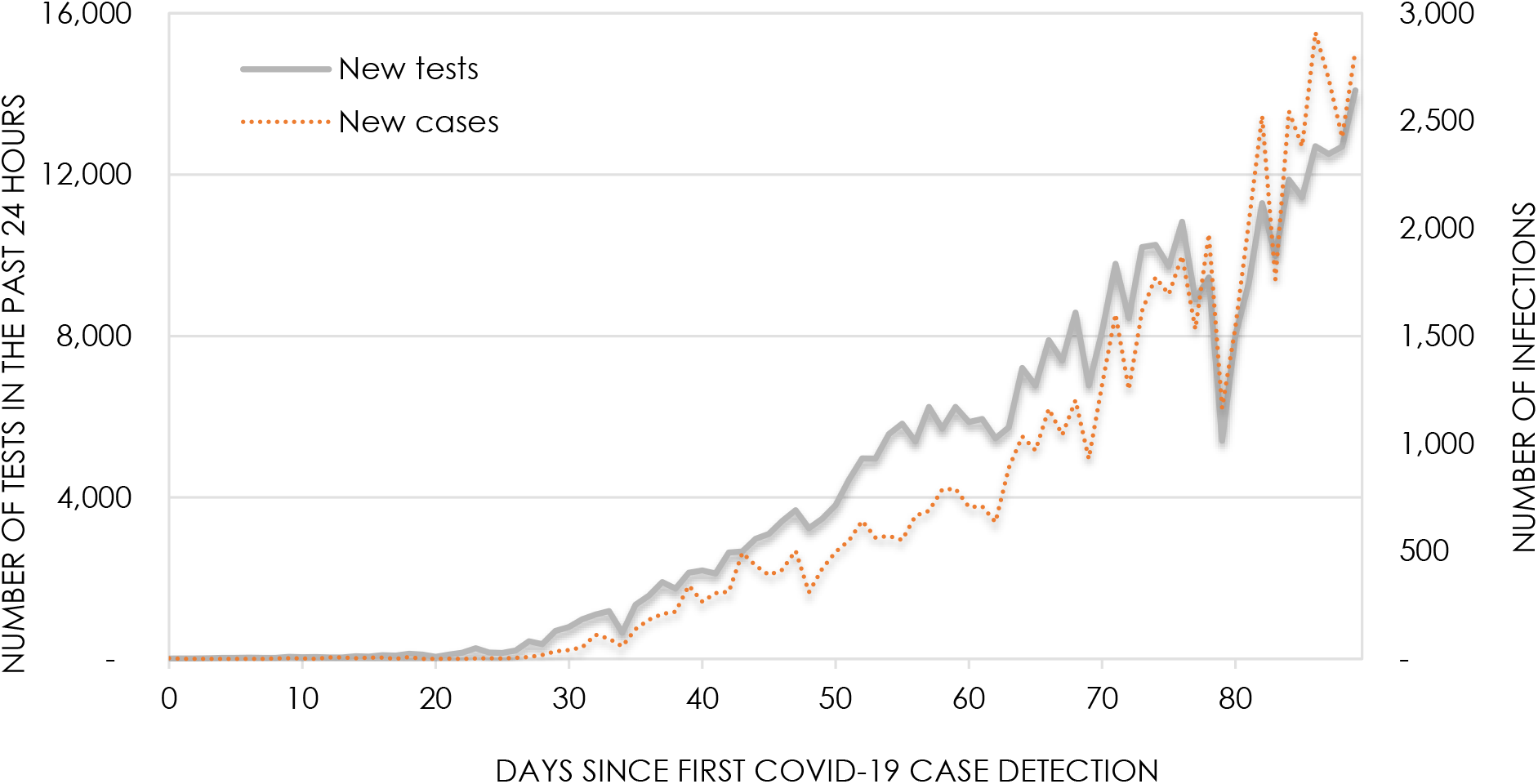
Trends in new Covid-19 cases and tests performed, Bangladesh 8 March–5 July 2020.

To decide on a sensible lag length for VAR, we ran lag-order selection diagnostics on both the variables. Since the diagnostic tests recommended four lags, we used that for VAR and applied small-sample corrections to the large sample statistics. As postestimation statistics, we performed Granger causality tests to assess VAR output. The test results show that the daily number of tests Granger-cause the number of new cases, and this findings is statistically significant (p <0.001) (See Table 1). We also tested whether the number of daily cases can Granger-cause the number of tests—since the p-value of 0.40 does not fall below the statistical significance threshold of 0.05, we couldn’t reject the null hypothesis that new cases do not affect daily tests performed.

**Table 1.** Granger causality test results.

| Dependent variable | Explanatory variable | F-statistic | Degrees of freedom (df) | Residual df | p-value |
| --- | --- | --- | --- | --- | --- |
| Daily new cases | Tests performed in the last 24 hours | 56.13 | 4 | 77 | 0.0000 |
| Tests performed in the last 24 hours | Daily new cases | 1.0184 | 4 | 77 | 0.4032 |

## Conclusion

An application of Granger causality test indicated that the number of daily new cases we observe for Bangladesh is perhaps due to an increase in test capacity rather than a change in the country’s Covid-19 infection rates. Unfortunately, the Granger causality test does not provide clear-cut results, and the results should, therefore, be interpreted with caution. From the results of this test we can infer that if the number of daily tests does not increase substantially, data on new infections will not give much information for analyses and projection.

Currently Covid-19 detection tests are being performed in 49 designated laboratories in Bangladesh, which requires immediate scale up to capture the actual disease progression in the country. In line with the recently published national preparedness and response plan for Covid-19 (GOB, 2020b), expanding laboratory network and testing capacity has to be seriously pursued. Apart from increasing the number testing facilities in the public sector, a public-private partnership can be explored to arrange Covid-19 testing in 354 registered ‘Category A’ diagnostic centers in the private sector (who provide microbiology and immunology test services to clients) (HSM, 2020). This would particularly be important to expand testing services at district-level, outside the large cities.

## Data Availability

Data are publicly available.

https://ourworldindata.org/coronavirus

## References

Government of Bangladesh (GOB). Corona Info. 2020a. [Accessed 12 July 2020] https://corona.gov.bd/.

Government of Bangladesh (GOB). National Preparedness and Response Plan for COVID-19, Bangladesh. Dhaka: Directorate General of Health Services. 2020b.

Hospital Services Management. Diagnostic Center Facility List. 2020. [Accessed 26 March 2020] http://103.247.238.81/hsmdghs/registration/hsm_facility_show_public.php.

Khan MHR. Covid-19 (Coronavirus) in Bangladesh, Report No. 63-08062020. 2020. [Accessed 12 July 2020]

https://sites.google.com/site/teachingsitehasinur/corona

Li Q, Guan X, Wu P, Wang X, Zhou L, Tong Y, et al. Early transmission dynamics in Wuhan, China, of novel coronavirus-infected pneumonia. N Engl J Med 2020; 382(13):1199–207; doi: http://dx.doi.org/10.1056/NEJMoa2001316.

Roser M, Ritchie H, Ortiz-Ospina E, Hasell J. Coronavirus Pandemic (COVID-19). 2020. [Accessed 08 July 2020] https://ourworldindata.org/coronavirus

World Health Organization (WHO). Director-General’s opening remarks at the media briefing on COVID-19. [Accessed 31 May 2020] https://www.who.int/dg/speeches/detail/who-director-general-s-opening-remarks-at-the-media-briefing-on-covid-1916-march-2020.

